# Health Provider and Sexual and Gender Minority Service User Perspectives on Provision of Mental Health Services During the Early Phase of the COVID-19 Pandemic in British Columbia, Canada

**DOI:** 10.1101/2022.02.18.22271151

**Authors:** A. M. Kennedy, S. Black, S. Watt, N. Vitkin, J. Young, R. Reeves, T. Salway

## Abstract

While the COVID-19 pandemic impacted everyone, social determinants of health and structural inequities have had compounding effects that shaped the experiences of some sub-populations during the pandemic. Stigmatization, discrimination, and exclusion contribute to a disproportionately high burden of mental health concerns among sexual minority (i.e., lesbian, gay, bisexual, queer, and other sexually-diverse) and gender minority people. Pre-pandemic, these health inequities are exacerbated by barriers to adequate mental health services including cost, waitlists, and experiences of sexual and gender minority stigma when accessing providers. During the COVID-19 pandemic, these barriers were further complicated by drastic changes in service delivery and access during the pandemic—i.e., a shift to online/virtual provision of care to reduce risk of COVID-19 transmission. To better understand the experiences of sexual and gender minority people accessing mental health services during the first three to nine months of the COVID-19 pandemic, we conducted semi-structured interviews with a purposive sample of 15 health care providers and administrators (summer 2020) and 14 sexual and gender minority individuals interested in accessing mental health services (fall 2020) in British Columbia, Canada. We used interpretive description to inductively analyze interview data. Triangulating between the provider and service user datasets, we examined changes in mental health and coping during the COVID-19 pandemic. We recorded increases in isolation and lack of identity affirmation; inequities in accessing mental health services during the pandemic, perceived opportunities for mental health support, and avenues for reducing mental health inequities through system-level changes that deserve particular attention during the pandemic.

## Introduction

The COVID-19 pandemic has significantly impacted social interaction, with a substantial reduction in in-person contacts (Ringa et al., 2021). These impacts are highly pronounced for populations burdened by social, economic, and structural inequities (Slemon & Jenkins, 2021; Rajkumar, 2020; Park et al., 2020; Jenkins et al., 2021; Holmes et al., 2020). Research suggests that public health measures designed to slow the transmission of COVID-19 have disproportionately affected the well-being of sexual and gender minorities (SGM), due to inequities in their participation in economic, political, and social systems, reduced social contact owing to real or perceived stigma, and increased isolation (Brennan et al., 2020; Gibb et al., 2020). Indeed, sexual minorities (i.e., lesbian, gay, bisexual, queer persons) and gender minorities face particular forms of oppression that may exacerbate the stressors of the pandemic, while also constraining opportunities for receiving support (Salerno et al., 2020). For example, social interaction, loneliness, and stress can have lasting effects on mental health outcomes. In this context, achieving a more fulsome understanding of the mental health of SGM during the pandemic will facilitate our comprehension of how to make mental health services more equitable, inclusive, and affirming (Gibb et al., 2020).

Between March 14 and December 12 of 2020, British Columbia (BC) underwent Phase 1, Phase 2, Phase 3a, and Phase 3b of its first reopening plan of the COVID-19 pandemic. In each phase, public health measures were adjusted according to COVID-19 cases (see Table 1 for the various phase restrictions).

**Table 1.** BC’s COVID-19 Reopening Phases and Restrictions. Informed by: Wadhwani, A. (2020, May 8). COVID-19: Here’s a phase-by-phase look at how B.C. hopes to re-open parts of society. *Victoria News. https://www.vicnews.com/news/covid-19-heres-a-phase-by-phase-look-at-how-b-c-hopes-to-re-open-parts-of-society/*

| Re-opening Phase | Dates | Restrictions |
| --- | --- | --- |
| **Phase 1** | March 14 - May 18 | • Entry of foreign nationals to BC banned<br>• Symptomatic individuals on flights to Canada banned<br>• Mass gatherings (>50 people) banned<br>• Restricted food and drink services<br>• US/Canada border closed to non-essential travel |
| **Phase 2** | May 19 – June 23 | • Start of BC’s reopening<br>• Health services, retailers, salons, in-person counselling, restaurants, museums, libraries, offices, sports, outdoor spaces, and childcare services reopened (with continued precautions)<br>• Students in kindergarten to grade 12 returned to school on a gradual and part-time basis |
| **Phase 3a** | June 24 – September 12 | • Non-essential travel in BC, hotels, TV/film reopen<br>• Return to in-class learning with partial weeks |
| **Phase 3b** | September 13 – End of study period (December) | • Starting the week of November 9, 2020, BC re-implemented their COVID-19 mitigation measures to interactions with one’s core bubble only |

The province defined ‘core bubble’ as “the people you spend the most time with and are physically close to. These would be people who are part of your regular routine, so household members, immediate family, a close friend or the people you have regular close contact with.” (Ministry of Health, 2020). In other jurisdictions that implemented similar public health restrictions (e.g., the UK), researchers noted that limiting contact to households or ‘bubbles’ has an inequitable impact on SGM, who are more likely to be living alone (Hafford-Letchfield et al., 2021). Knowing the role of stress and isolation in the context of these pandemic mitigation measures is important to bolster support for sexual minorities as the COVID-19 pandemic progresses.

Jenkins et al. (2021) noted that 45% of SGM surveyed reported worsened mental health during the first few months of the pandemic, as compared with 38% of cisgender/heterosexual respondents. Pre-pandemic research helps to explain these findings by identifying that mental health disparities among SGM are attributable to multiple factors. These are: cumulative personal histories of discrimination, stigma, insensitivity, microaggressions, negative health care experiences (e.g., denial of treatment, endorsement of heteronormative culture, heterosexist terminology), and structural inequities (e.g., limited employment or housing opportunities) related to sexual orientation-based discrimination (Gonzales et al., Hatzenbuehler, Phelan, & Link, 2013; Nock et al., 2008; Salway et al., 2019; Marshal et al., 2011; Hottes et al., 2016; Salway & Gesink, 2018; Plöderl et al., 2018; Hammack et al., 2017; Hatzenbuehler et al., 2010; Ferlatte et al., 2015). This is clarified when looking at the Minority Stress Model (Brooks, 1981), which posits that biophysical and psychological health are impacted by cultural and social stressors, including experiences of discrimination, exclusion, and marginalization among SGM. SGM in turn face a disproportionate amount of stress related to isolation and social disconnection (Rich et al., 2020). These factors became more prevalent and frequent during the early phase of the COVID-19 pandemic when physical distancing measures were enacted and non-essential businesses closed (Rumas et al., 2021). Jenkins et al. (2021) found that 37% of surveyed SGM reported experiencing loneliness or isolation (compared to 30% of non-SGM participants), and that 65% were experiencing separation from family and friends during the pandemic (compared to 59% of non-SGM respondents) (Jenkins et al., 2021).

SGM also face unique barriers to accessing mental health care, including avoidance of social interaction with health professionals due to fear of experiencing additional minority stress during these interactions (Salway et al., 2018). Therefore, providing referrals to affirming support is important. Given that sexual health services fill important gaps in health care opportunities for SGM who do not have other primary or mental health care (Salway et al., 2019; Salway et al., 2021), it is also important to include these settings when developing a holistic understanding of sexual/mental health care access. Therefore, providers from sexual health clinics are included in this analysis, given that they often provide care to individuals who face barriers accessing a primary care provider, or who are deterred from other primary care settings due to privacy concerns, stigma, and/or embarrassment (Black et al., 2020). Additionally, mental health services can be complicated to navigate, and many individuals who may benefit from accessing this kind of support do not seek it. As BC shifted to online mental health care provision during the COVID-19 pandemic to reduce the risk of COVID-19 transmission (CBC News, 2021), this may have exacerbated access barriers for some SGM. To investigate these barriers and the mental health needs of SGM populations in BC during the early stages of the COVID-19 pandemic, we interviewed health professionals with extensive experience in the health of SGM, as well as SGM sexual health service users.

## Methods

### Sample

We used two different samples to triangulate between the perspectives and experiences of SGM accessing services (hereafter ‘service users’) and health professionals with extensive experience serving SGM clients (hereafter ‘providers’). Between May and September of 2020, we recruited and interviewed a purposive sample of 15 health care providers and administrators working with SGM in BC (see Table 2). We used snowball sampling to recruit these individuals, and shared letters of invitation through the <u>2S/LGBTQ+ Roundtable</u> and the BC Sexually Transmitted Infection/HIV Provincial Nurses Working Group.

**Table 2.** Characteristics of Provider participants included in analysis.

|  |  |
| --- | --- |
| Participants | 15 |
| **Type of Medical Setting<sup>a</sup>** |  |
| *Sexual Health Clinic/Organization* | **6** (40.0%) |
| *Primary Care* | **3** (20.0%) |
| *Outreach Care* | **2** (13.3%) |
| *Youth Clinic* | **1** (6.7%) |
| *Private Practice* | **2** (13.3%) |
| *Health Authority/Regional Public Health Organizations* | **3** (20.0%) |
| *Community Health* | **2** (13.3%) |
| *Not-for-profit Mental Health* | **1** (6.7%) |
| *Queer Health Organization* | **3** (20.0%) |
| **Geography** |  |
| *Greater Vancouver* | **12** (80.0%) |
| *Other Parts of the Province* | **3** (20.0%) |
| **Profession<sup>b</sup>** |  |
| *Nurse* | **6** (40.0%) |
| *Nurse Practitioner* | **1** (6.7%) |
| *Mental Health Provider* | **6** (40.0%) |
| *Health Promotion Case Manager* | **3** (20.0%) |
| *Health Educator* | **4** (26.7%) |

<sup>a</sup>One participant may have more than one setting.
<sup>b</sup>One participant may have more than one profession.

In the fall of 2020, our team also interviewed a purposive sample of service users who were respondents to the “Sex in the time of COVID-19” survey (Gilbert et al., 2021), and indicated interest in being involved in our study. To be eligible to complete that survey participants had to meet the following criteria: 1) existing clients of BC Centre for Disease Control (BCCDC) sexually transmitted infection (STI) clinics or Get Checked Online, 2) be at least 16 years old, 3) have either visited the BCCDC STI clinic or been tested in the previous year (March 15, 2019 – March 17, 2020). From this purposive sample, we interviewed and surveyed 27 individuals. For the analysis in this paper, we have only selected the participants who self-identified as gender or sexual minorities in the survey (n=14) (see Table 3).

**Table 3.** Characteristics of Service Users included in analysis.

|  |  |
| --- | --- |
| Participants (meeting criteria) | 14 |
| **Age (mean, range)** | **(20-54 years)** |
| **Age Category** |  |
| *20-29* | **5** (35.7%) |
| *30-39* | **8** (57.1%) |
| *40-49* | **0** (0.0%) |
| *50-59* | **1** (7.1%) |
| **Gender Identity<sup>a</sup>** |  |
| *Woman* | **5** (35.7%) |
| *Man* | **8** (57.1%) |
| *Non-binary* | **1** (7.1%) |
| *Genderfluid* | **1** (7.1%) |
| **Transgender** |  |
| *Yes* | **0** (0.0%) |
| *No* | **14** (100.0%) |
| **Sexual Orientation<sup>b</sup>** |  |
| *Bisexual* | **6** (42.9%) |
| *Gay* | **6** (42.9%) |
| *Pansexual* | **1** (7.1%) |
| *Mostly Straight* | **2** (7.1%) |
| **Race/ethnicity** |  |
| *Chinese* | **1** (7.1%) |
| *Iranian* | **1** (7.1%) |
| *Multi-racial* | **1** (7.1%) |
| *White* | **11** (78.6%) |
| **Health Authority Participants Currently Living in** |  |
| *Vancouver Coastal Health* | **7** (50.0%) |
| *Interior Health* | **3** (21.4%) |
| *Island Health* | **4** (28.6%) |
| <sup>a</sup>One participant selected more than one gender identity. |  |
| <sup>b</sup>Participants were able to describe sexuality in their own terms. |  |

### Data Collection

Semi-structured interviews with mental and sexual health providers and administrators were conducted over Zoom by four members of our team during the summer (May-July) of 2020. The interview guide included four main parts: 1) Interviewee’s role and experience as a service provider, 2) How the pandemic affected their service users, 3) Barriers services users experienced when trying to access mental health services, including COVID-19-specific barriers, and 4) Knowledge and use of existing mental health and substance use resources. Parts 2 and 3 are the focus of this article. Questions within these broader themes were answered by service providers specifically, as they were able to draw on their professional mental health experience and trends noticed across multiple service users.

The interviews with service users, conducted between October-December of 2020, covered a range of topics related to mental health needs of sexual health service clients and desires for sexual/mental health service integration. More specifically, the interviewees were asked: (1) How has the pandemic affected you? (2) What changes have you noticed in your mental health during the COVID-19 pandemic, if any? and (3) In what ways has the COVID-19 pandemic affected your access to mental health services? Participants were asked to complete a demographic questionnaire after the interviews. The gender and sexual orientation of participants identified throughout the manuscript is informed from the responses to this questionnaire.

All interviews were conducted over Zoom and lasted roughly one hour. With participant permission, interviews were recorded and saved to a secure database. All recordings were externally transcribed, and then internally accuracy-checked by members of our team who did not conduct the interview. This study was reviewed and approved by the Simon Fraser University Research Ethics Board, and the UBC Behavioural Research Ethics Board.

### Analysis

Our team conducted an inductive thematic analysis in NVivo 12. When conducting inductive thematic analysis, researchers searched for emerging themes in the data which aided in describing the phenomena of focus, and which then became the categories of analysis (Braun & Clarke, 2006). Thematic analysis occurred in phases, including becoming familiar with the data through transcript cleaning, generating initial codes, categorizing codes into potential themes, checking these codes for value in multiple transcripts, consistent and ongoing defining and naming of each theme, and the selection of illustrative quotes and report writing (Braun & Clarke, 2006). To uphold rigor in our coding process, two individuals from our research team coded each transcript (Saldaña, 2015). Quotations were accompanied by type of workplace setting and profession, in the case of service providers, and self-described gender and sexual orientation identities, in the case of service users.

The two datasets were analyzed separately, yet similar themes arose related to the COVID-19 pandemic and mental health in both datasets. Thus, in order to illustrate synergies between study population experiences, results were presented together under shared themes. Triangulating (Heale & Forbes, 2013) between the two datasets, our team was able to examine perceived barriers to and opportunities for sexual/mental health care in the context of physical distancing. Pulling from provider and service user perspectives allowed us to determine avenues for reducing mental health inequities through system-level changes that deserve particular attention during the pandemic.

## Results

Our findings were presented in two parts. First, we interpreted interviewees’ responses to how the mental health of SGM has been affected by the pandemic, with particular focus on increased isolation, and changes to mental health. These emerged as prominent and recurrent themes in our data, which in part can be explained because of the role minority stress has in the health and wellbeing of SGM (Brooks, 1981). Second, we investigated barriers to accessing mental health services that were exacerbated during the pandemic, including the perceived barriers to sexual/mental health care related to physical distancing, privacy and safety considerations, and other inequities to accessing affirming sexual/mental health care.

### Direct Mental Health Effects of Pandemic-related Measures

#### Changes in Mental Health and Coping over Time

Both service users and health providers described dynamic experiences and mental wellbeing that changed at various stages of the pandemic. Service users specifically shared how the first few months of the pandemic were often less stressful than normal, due to less external pressures (e.g., work, social gatherings), freeing up time for working on their mental health. However, they specified that ongoing restrictions in the later months of the pandemic, coupled with worry about the future, negatively affected their wellbeing and coping mechanisms:

> …for the first few months of [the pandemic], [my mental health] was actually better. It was weird ‘cause I hadn’t had depressive episodes or a lot of anxiety because [I had] nothing to do, so I was really chilling for a couple of months there. And then it wasn’t until you had to really get back into life with the [COVID-19] exceptions and everything that it kind of started stressing me out a little bit more and I got into my normal habits of, you know, mental health [concerns]… (Service user, female, bisexual)

Some health providers identified that some SGM encountered less stigma because they were spending more time at home:

> …some of the clients I’ve worked [with] before have said they have to do less work navigating the world and people’s reactions to them, they have to do less self-advocacy because they’ve just been at home and that’s actually been really positive… (Nurse, interviewed phase 3)

Some service users supported this sentiment and shared that the pandemic took social and work pressures out of their lives and provided them with more time to engage in self-reflection, reach out for mental health support, and prioritize self-healing. One service user shared this view of experiencing both concern and relief by saying: “I can finally breathe, but the world is on fire” (Service user, genderfluid/female, straight-ish) – indicating the complexity of consequences and opportunities in the pandemic.

Providers similarly identified changes in individual access to mental health support. In the early phases of the pandemic, providers shared:

> …our practice has really slowed down… and I don’t know what the reason for that is, I don’t know if it’s because people don’t have money [and] they’re not working. …But across the board we have noticed a drop in free and paid counselling [and] requests for free and paid counselling… (Mental health provider, interviewed in phase 1)

As the pandemic persisted, some mental health providers noted that individuals started accessing services again: “…a lot of colleagues also mentioned a drop-off in new clients since the pandemic, and that’s starting to resume” (Mental health provider, interviewed in phase 2). Providers cited several possible reasons for this trend in service uptake, including an initial lack of knowledge about which services were operating:

> …people kind of thought that services weren’t being offered but they saw the whole economy shut down and just assumed that we would be doing the same and of course we did adapt, like going virtual but that wasn’t always being considered by potential clients… (Health promotion case manager, interviewed phase 3)

They also attributed a lack of confidence in remote mental health support to these shifting trends:

> I do the Zoom counselling now and I love it, but we’ve also had a lot of clients who have totally disengaged with their counsellors because even though they could do it and they have access, they don’t want to do it… (Nurse and health educator, interviewed in phase 3)

#### Isolation and Identity Erasure

Service users and providers identified that during the pandemic, mandates for physical distancing and self-isolation worsened exclusion, social isolation and reduced social support for various marginalized populations, exacerbating mental health concerns. One provider alluded to this by saying:

> …a lot of queer guys… have mental health needs … right now especially those who maybe don’t have nuclear families. You know there’s … a number of cultural reasons gay men …may live alone for the rest of their lives and you know their friend circle is one that they are really reliant on for the supports that they would typically require and because of, especially early in the pandemic, the strictness of social distancing, and whatever else, they’re really missing those supports and …I know it’s something that we all sort of experience but it may be heightened in those individuals. (Health promotion case manager and health educator, interviewed phase 3)

Several service users addressed this phenomenon when they expressed the negative mental health effects of being distanced from their groups of friends during the pandemic:

> …I do feel that [the pandemic has] affected me mentally just from a lot more isolation with myself and just a lot, you know I actually I like my own company, it’s not that I’ve never lived alone…but just having that kind of connection with somebody that you, you know I have my bubble and my friends that I’m with but I don’t know it’s just, it feels a little bit more isolated… (Service user, male, gay, interviewed in phase 3)

Given that social conditions are a recognized fundamental determinant of disease (Marmot & Wilkinson, 2005), the negative health effects of social isolation are of immense concern during the pandemic, particularly among individuals who may rely on the connection of individuals outside of their current household.

Additionally, some sexual minorities experienced drastic restrictions in their ability to fully express their sexual orientation during the pandemic. This was addressed by a clinical counsellor when they said:

> I…[work] with non-monosexual folks, so meaning people who are attracted to folks of more than one gender and I know in this time, for example, a lot of people who identify as bisexual who are live-in partners with folks of one gender may be experiencing sort of what they sometimes term as a re-closeting or just not having access to the other identity affirming activities that they incorporate into their own sort of structures for identity affirmation. (Mental health provider, interviewed phase 1)

This “re-closeting” can be damaging to the identity affirmation, and subsequent mental wellbeing, of SGM. Similarly, service users experienced the negative impacts of relationship structure changes:

> […] we are polyamorous, but we decided to close up the marriage because of COVID too so that’s just like a new big change. So yeah we are at home a lot and I do think it’s making me a little like socially more challenged, you know like nervous for when it goes back to normal, that we’re all going to be weird. (Service user, female, bisexual, interviewed in phase 3)

Another way that SGM experienced social isolation during the pandemic was in the closure of in-person queer leisure spaces, pride celebrations, and queer community events (Brennan et al., 2020). This is particularly concerning as these spaces and celebrations have historically been used by SGM to “provide safe and appropriate contexts for the expression of [queer] identities” (Anderson & Knee, 2020). One provider shared:

> [For] a lot of folks you know there isn’t that sense of community where they live, so they go say to the [gay neighborhood] to gay bars to connect with other folks, or to bathhouses. But they can’t do that anymore, so they feel much more isolated. They’re tending to, you know, self-medicate to cope with their feelings which further causes them to withdraw a bit more… (Health promotion case manager, interviewed phase 2)

These restrictive social connection changes and the shift to isolation may also have gone against a person’s personality and led to feelings of alienation: “I’m a very social person and I hate that I’m isolated all the time, and even this [interview] is great ‘cause it’s interaction” (Service user, female, bisexual). One provider alluded to this trend when sharing that this distance from community can remove feelings of safety:

> …I think the pandemic has really, yah it’s hard, we rely on our people to create these bubbles of safety - to remind us that we exist, to remind us that we are wonderful and beautiful, and we can’t meet with them as easily so that’s kind of hard. (Mental health provider, interviewed phase 1)

Distance from these networks was not only detrimental to mental wellbeing, but also damaging to some SGM’s identity expression. \

### Inequities in Accessing Mental Health Services During the Pandemic

While some mental health services shifted to a remote format, several concerns came up around the need for a safe and private space to access online mental health services. While these themes did not come up in the service user interviews, several of the health providers we interviewed cited their concern that individuals who need support may not be able to access it, and that accessing online mental health services when the conversations may be overheard could be dangerous for some individuals who may experience intersecting stigma about both accessing mental health services and talking about sexual identity. One service provider shared with us that:

> …[their] biggest fear right now is about how many folks are at home especially folks with marginalized gender and sexual identities that are not in safe spaces for them and don’t… have access to any privacy that would allow them to even reach out for the mental health support that they need. (Mental health provider, interviewed phase 1)

This highlights an important consideration regarding mental health service access during the pandemic: SGM may have been living with others not affirming of their identities, thereby increasing their need to access mental health support. However, those same living situations may have been a barrier to reaching out to providers. This concept of intersecting safety concerns and reduced care access also came up as a consideration for SGM experiencing interpersonal violence during the pandemic:

> …counsellors providing care over the phone or by video … have to be much more hesitant to work with people who are at risk of interpersonal violence, because there’s a higher chance that if a partner finds out there may be increased violence… So it’s quite concerning to think about the fact that the [COVID-19] context may be increasing risk and significantly decreasing access to care. (Mental health provider, interviewed phase 1)

This demonstrates how privacy and safety inequities experienced during the pandemic may have been worsened by the change in service format delivery to remote services (Richardson et al., 2020).

Several other inequities also materialized around having the technological and financial means to access online services. One provider said:

> …it’s definitely right now where who I see are people who can afford to pay private practice fees - who are employed, who may or may not have benefits, extended health insurance. So I’m definitely seeing a more [economically] privileged segment of the queer community, and having worked for the first half of my career in non-profits, like I really recognize the privilege, who I get to see and the kind of work and conversations I get to have now. (Mental health provider, interviewed phase 1)

This indicates whose voices are being heard, who is getting support, and who is experiencing challenges in accessing mental health services. Specifically, SGM who were economically disadvantaged may have had particular barriers to accessing remote mental health care.

### Perceived Opportunities for Remote Sexual/Mental Health care

It is evident from previous research that the COVID-19 pandemic and the shift to remote mental health services had a negative impact on SGM’s mental wellbeing and access to sexual/mental health care. However, our findings from both sets of interviews show that, in some cases, remote services and physical distancing requirements created new opportunities for increased accessibility to mental health support. More specifically, several providers shared how facets of the shift to remote mental health service delivery had some positive implications on health equity, including acceptance of remote mental health support:

> …[the pandemic] made [remote] counselling a lot more acceptable and so that will hopefully allow… different organizations and different workers to bridge those gaps in the geography. (Health promotion case manager and health educator, interviewed phase 3)

With this institutional shift in acceptance of remote mental health services, mental health care has been made more accessible to individuals living in rural regions or those who are unable to access in-person services for various reasons (e.g., chronic pain or fatigue). Remote mental health services can also provide increased accessibility for individuals who may experience stigma associated with accessing mental health support. One service user spoke to the challenges of accessing face-to-face mental health services in rural communities:

> …when it comes to face-to-face like I couldn’t imagine what that might be like in more so rural communities…being from those small communities and the stigma behind accessing those services. I think it is like changing that, like we are a bit more accepting of accessing, people accessing those things ‘cause I think everyone should. It’s yeah so it could be that kind of small community, small-minded kind of thing where you’re like “I don’t want to be known as the crazy person” or anything like that or kind of thing. (Service user, genderfluid/woman, straight-ish, interviewed in phase 3)

Remote mental health services also created more equitable access for some SGM who may not have had access to a space specifically designed for them, or who may not have felt safe going into a non-affirming mental health space due to stigma from other service users, providers, or both:

> … [mental health services] going virtual because of COVID … [is] a big silver lining, especially when we’re trying to reach a population that for a number of reasons may not have access, or may not be safe going into a space that’s specifically queer, or who live in places where such services just aren’t available to them. (Health promotion case manager and health educator, interviewed phase 3)

Therefore, remote services can provide care to SGM who don’t have local affirming providers by working with providers who are not geographically close to them.

### System-Level Needs for Equitable Mental Health

We identified several themes pointing to opportunities to improve mental health care access and support in both datasets. The first is the call for more publicly funded/accessible mental health services. This is particularly important given that mental health care is not covered in basic health coverage in Canada, and that many employee benefit programs limit the coverage of mental health care. A lack of options places financial pressure on SGM to access non-affirming care. SGM-affirming mental health support includes various aspects (e.g., conversational, environmental, social) aimed at helping SGM feel validated, valued, and respected. Therefore, SGM accessing non-affirming care are at risk of experiencing stigmatizing interactions, or spending more time educating the provider than receiving care themselves:

> Having more consistency between employers on how much coverage they get for counselling [would be helpful]. Like for some people they don’t get any coverage, they

just get the employee assistance program, which in some ways is limited and tends to be not so helpful especially if you’re gay or needing a gay-affirming counsellor. There’s a number of clients who I’ve have talked who have said, ‘yah I tried a few EAP counsellors but I spent more time trying to explain to them what it was like being gay, and the culture around being gay, or having an open relationship, or engaging in like fetish sex or kink sex and it was like I was doing more of the educating than the therapist’, and that was a waste of their time in some ways as opposed to having like someone knowledgeable, more informed about the community and the culture, and then being able to not have their ignorance be the topic of therapy. (Mental health provider, interviewed phase 2)

Similarly, one service user shared the importance of seeing a mental health provider who identifies as a sexual minority:

> …I specifically went [to a health service for gay men] because I’m a gay male, and … to have a guy that’s potentially had the same experiences, you know understanding my desires you know or my sexual preferences, and I get that there’s no judgement but it’s just more, I feel that there’s just a bit of a better understanding and a connection. (Service user, male, gay, interviewed in phase 3)

Since SGM may face financial barriers to affirming mental health support, it is particularly pressing to make affirming mental health care more accessible.

Another core suggestion was to decrease social isolation by bolstering programming focused on helping SGM build relationships and networks. Both service users and providers described pre-pandemic activities and programs that were successful in connecting SGM to their community:

> …there is you know a lot of gay guys here, it’s huge the population and I didn’t really realize how many there was, to be honest when I first moved here…I do feel that the community would be better with some form of … [activities to facilitate] connection … I’ve kind of been to, or seen, really good examples actually of at the [local pub] on the Wednesday before COVID like the gay men’s choir they go in there after their rehearsals and it’s so much fun… Things like that are really supportive to people, and it really makes people get connections for sure. (Service user, male, gay, interviewed in phase 3)

Service providers also shared the impression that connection to community and social networks is important for the mental health of SGM. Programming was of particular importance in proactively addressing social isolation during the pandemic while individuals were feeling heightened isolation:

> …[We need to figure] out how to create programs that are decreasing social isolation, but not only for folks who want to talk about their problems, like for folks who just want interaction and thinking of that as a part of the mental health mandate. (Mental health provider, interviewed phase 1)

Particularly significant here is not only bolstering inclusive and affirming programming, but also integrating proactive methods to reduce social isolation. This should be viewed as part of the mental health mandate, rather than a supplementary option.

## Discussion

This study explored how the COVID-19 pandemic affected SGM’s mental health and access to mental health support. Our findings indicate that isolation due to physical distancing requirements is one of the most pressing concerns for SGM during the pandemic. Additionally, while some SGM experienced worsened mental health and service access in light of remote mental health care provisions, others simultaneously experienced improvements in access, suggesting a need to better understand which SGM are served by digital health interventions, and why.

Epidemiological studies of the (predominantly heterosexual) general population pointed to a return to baseline mental health status (regarding anxiety, depression, distress, loneliness) by mid-2020 (Aknin et al., 2021; Luchetti et al., 2020); however, public health interventions that do not account for differential exposures to risks and resources across socially defined subpopulations may miss within-population disparities and trends (Frohlich & Potvin, 2008). Indeed, the impact of the pandemic has not been evenly distributed across the population (Saban et al., 2021; Jenkins et al., 2021). There is good reason to prioritize tailoring our public health response to sub-populations that have historically faced barriers to equitable service provision (Jenkins et al., 2021).

Although mental health inequities existed for SGM before the pandemic, various social and mental health inequities have been exacerbated during the COVID-19 pandemic due to the provincial mandates to physical distancing orders and service closures. Our research aligns with the existing body of literature, which has identified that the mandates for physical distancing and self-isolation have contributed to particular forms of exclusion, social isolation and reduced social support for SGM (Daly et al., 2021; Kneale & Bécares, 2021). The COVID-19 pandemic is particularly concerning for SGM who may have recent and acute experiences of minority stress, and may also be experiencing isolation and separation from affirming persons, resources, social outlets, and supports (Brennan et al., 2020). This can be the result of the closures of affirming activities and returning to living with families of origin (Salerno et al., 2020). Since chosen family (i.e., family groups determined by choice rather than biological relationship) and support networks are of great importance to SGM, a separation from these networks within rigid or misinterpreted physical distancing public health measures can be greatly damaging (Moore et al., 2021). During the COVID-19 pandemic, some SGM lived in unsafe spaces, such as living with unsupportive families leading to increased risk of violence and/or abuse (Gibb et al., 2020). These experiences can amplify various and long-lasting physical and mental health concerns (Gibb et al., 2020).

For some, the shift to remote mental health support increased service accessibility, and for others, this created barriers in access. Some sexual and gender minority individuals – particularly those in rural/remote settings, physically unable to attend in-person mental health support sessions, and who feel anxiety upon physically entering spaces that may not be affirming of SGM – experienced more accessibility to mental health support through remote mental health services. However, for SGM who did not have access to affirming or safe/private spaces at home (e.g., those living with families of origin who are not SGM-affirming), accessing remote mental health support was not a safe option (Fish et al., 2020). Access inequities can also result from shifts in physical location during the pandemic, like moving in with family members, or leaving university campuses; all of which can result in re-closeting concerns (Fish et al., 2020). This change in location can sever people’s connections to their mental health providers if location-based restrictions and conditions exist for a provider (e.g., condition that the service user must reside in a particular geographic area to be eligible to access the service). It can also worsen general mental health if the change in location is to a non-affirming environment, community, or living situation. Beyond service provision, providers also addressed how intersecting factors like socio-economic status can impact people’s experiences and mental health during the pandemic, including concerns about employment stability, expenses, and ability to access services and other recreational activities. These considerations influenced whom they were seeing in their remote mental health sessions (i.e., individuals who could afford mental health support).

Our findings alluded to a series of system-level recommendations for policies, programming, and support for SGM. Specifically, we heard the need for low-cost counselling and SGM-affirming mental health support groups, since these services can often be expensive, with long waitlists and onerous intake processes. Our participants (providers and service users) identified the importance of bolstering widespread support and advocacy efforts towards system-level changes and improvements to mental health equity for SGM – not only during the pandemic, but into the future, as well as the need to expand affirming conversations about mental health and bolstering mental health equity for those individuals who experienced challenging access during the pandemic.

## Conclusion

Triangulating between provider and service user datasets, we examined the perceived barriers to and opportunities for sexual/mental health care during the COVID-19 pandemic among SGM in British Columbia, Canada. While we found that the pandemic exacerbated existing inequities, it also gave rise to some equity-centred mental health opportunities, including (1) providing the option for virtual counselling; (2) creating more opportunities for support for individuals living in rural and remote communities, as well as others who are unable to access services in-person; and (3) reducing potentially stigmatizing person-to-person interactions. Participants emphasized the need for increasing publicly funded/accessible and SGM-affirming mental health services; expanding SGM-affirming counselling ; and decreasing social isolation by bolstering programming focused on providing spaces for SGM community building and connection. Future research should also focus on expanding what it means to provide affirming mental health care.

## Data Availability

Data are not available owing to privacy concerns.

